# Costly Miscommunication: College Students with Disabilities’ Perceptions of COVID-19 Vaccine Costs

**DOI:** 10.1101/2021.11.04.21265924

**Authors:** Z.W. Taylor, Chelseaia Charran

## Abstract

Institutions of higher education have mandated COVID-19 vaccinations for students wishing to return to an on-campus, in-person learning experience. However, college students with disabilities (SWDs) may be hesitant to take a COVID-19 vaccine for a variety of reasons, possibly delaying or denying these students’ access to higher education. Yet, an under-researched aspect of COVID-19 vaccinations and related communication is whether college students with disabilities understand that the COVID-19 vaccine is free and whether that understanding varies by intersectional identities. As a result, this study’s research team surveyed 245 college students with disabilities to explore these students’ knowledge of vaccine costs and whether differences exist between groups. Data suggests many college students with disabilities do not know that COVID-19 vaccinations are free: White/Caucasian SWDs were most aware of COVID-19 vaccines being free (23.6%), while Latinx students were least aware (1.3%). Moreover, women were more aware of free COVID-19 vaccines (14.8%) than men (11.4%), first generation college students were more aware (15.6%) than non-first generation college students (12.2%), and full-time students (19%) were more aware than part-time students (8.9%). Overall, less than 25% of SWDs understood that COVID-19 vaccines are free. Implications for health communication, vaccine awareness, and higher education policy are addressed.

## Costly Miscommunication

### College Students with Disabilities’ Perceptions of COVID-19 Vaccine Costs

The COVID-19 pandemic has fundamentally changed how United States (U.S.) institutions of higher education operate, including a wealth of additional safety protocols to ensure that if students, faculty, staff, and other stakeholders are on campus, these individuals can remain safe (Gonzalez-Ramirez et al., 2021; Lederman, 2020). Catalyzing the ability for campuses to re-open their doors has been the rapid development of COVID-19 vaccines, including many U.S. institutions feeling so confident about vaccine efficacy that they have mandated students, faculty and/or staff be vaccinated before returning to campus and taking classes in person or living in residence halls (Nguyen et al., 2021; Redden, 2021).

Even though many U.S. institutions feel confident in COVID-19 vaccines, many postsecondary students of traditional college-going age (ages 18-24) have expressed skepticism and hesitancy to be vaccinated of a concern for their health and the health of others. In late April 2021, a Harris Poll found that 11% of Generation Z will not get the vaccine and 31% assert that they will wait an indeterminate amount of time before considering getting a vaccine (Owens, 2021). The same poll found that women, people of color, those living in rural areas, and people with disabilities are least likely to get the vaccine, while men, Whites, and those living in urban areas expressed the greatest likelihood of getting the vaccine. Yet to date, little is known about how college students with disabilities—an extremely marginalized population in U.S. higher education—may express vaccine hesitancy and what can be done to assuage concerns and encourage vaccination.

Prior research into vaccine hesitancy often contributes to failed vaccination campaigns, and thus, and continued spread of preventable infectious diseases (Dubé et al., 2013). The causes behind vaccine hesitancy are multifaceted, with studies explaining that parents of children with disabilities often express high levels of vaccine hesitancy due to their child’s health and the anxiety surrounding the potential effects of the vaccine (Salmon et al., 2015). Moreover, many parents are susceptible to the *post hoc ergo propter hoc* logical fallacy, or *after this, therefore because of this* as it relates to vaccines, often falsely believing that a child’s illness and/or disability is due to vaccines, and therefore, all vaccines are inefficacious or unsafe (Salmon et al., 2013). Salmon et al.’s (2013) study outlined several categories of vaccine hesitancy, including the natural versus *manmade hesitancy* (diseases are natural but vaccines are manmade and thus dangerous), the *predictable hesitancy* (disease impacts are known but vaccine impacts may not be known), and the *not dreaded* hesitancy (some diseases are not dreaded but adverse vaccine side effects are dreaded), many of which may be experienced by modern college students with disabilities.

What is rarely considered and analyzed by researchers is vaccine cost as it influences vaccine hesitancy. In an era of misinformation, fake news, and politicized reporting, a March 2021 U.S. Census survey found that nearly 90 million U.S. residents were hesitant to take a COVID-19 vaccine and 7 million of those residents expressed hesitancy over cost concerns (United States Census Bureau, 2021). These findings are problematic, as the United States Federal Government and the Centers for Disease Control and Prevention (2021) have informed the U.S. public that the “federal government is providing the vaccine free of charge to all people living in the United States, regardless of their immigration or health insurance status” (Centers for Disease Control and Prevention, 2021, para. 33), and that all COVID-19 vaccination providers cannot “Charge you for the vaccine” or “Charge you any administration fees, copays, or coinsurance” (Centers for Disease Control and Prevention, 2021, paras. 35-36). In U.S. contexts, vaccine hesitancy over cost may be misguided, as COVID-19 vaccines will be free for the foreseeable future.

Related to young adults, specifically college students with disabilities, no extant research has explored what college students with disabilities know about the COVID-19 vaccine and whether these students understand that the COVID-19 vaccine is free. It is critical to gauge the knowledge level of college students in this area, as many of these students may delay their education, and thus, their improved job prospects and financial standing if they are vaccine-hesitant for an illogical reason. Moreover, countless institutions of higher education will require COVID-19 vaccines to return to in-person on-campus learning, yet college students may not re-engage themselves with their institution if they view a COVID-19 vaccine as unaffordable, stigmatizing low-income students with disabilities. As a result, this study surveyed 237 current college students (enrolled as of March 2021) to answer these questions related to COVID-19 cost and vaccine hesitancy among college students:

1. Do college students with disabilities understand the COVID-19 vaccine is free?
2. Does knowledge of vaccine cost vary by identity (race, gender, first-generation in college status, etc.)?

Answering these questions will inform both the scientific and educational community regarding how to promote COVID-19 information especially as it relates to cost, as the United States attempts to achieve herd immunity and promote COVID-19 vaccination efforts across the country.

## Methods

Data for this survey were gathered in March 2021 when public availability of COVID-19 vaccines in the U.S. became clearer through official communication from the Centers for Disease Control and the World Health Organization (Centers for Disease Control and Prevention, 2021). The research team employed Amazon Mechanical Turk (MTurk) to survey postsecondary students currently enrolled at institutions of higher education. MTurk has been found to be a unique and robust source of human intelligence services, including survey completion in educational contexts (Follmer et al., 2017). Several recent studies in education focused on financial aid jargon (Taylor & Bicak, 2019) and computer science education (Hellas et al., 2020) have used MTurk to answer research questions that require a large, nationally representative dataset, akin to the study at hand related to college students’ with disabilities attitudes toward COVID-19 vaccinations.

The survey first asked people if they were currently enrolled college students, and then asked for students to self-identify as having a disability. If a person answered either question with a ‘no,’ the person was not allowed to finish the survey. The survey asked for a student’s birth year, race (Asian American/Pacific Islander, Black/African American, Latinx/Hispanic, White/Caucasian, or a fill-in-the-blank), gender (woman, man, non-binary conforming), first generation in college status (defined as neither parent earning any level of postsecondary credential), educational level (two-year, four-year, or graduate), enrollment status (part- or full-time), and current mode of education (on-campus, online, or hybrid). Then, students were asked one question:

1. How much will the COVID-19 vaccine cost you? (Nothing - the vaccine will be free through insurance or the federal government/$1-99/$100-199/$200+)

## Limitations

As with any survey study, this study is limited primarily by the reliability and validity of the survey data. This study gathered data from MTurk and participants who self-reported their college enrollment status, disability status, as well as other demographics. Moreover, this study is also limited by its temporal nature, meaning that attitudes toward COVID-19 vaccines may drastically change over time as vaccine efficacy is reported and vaccines become safer and more available to the general public. In addition, as institutions of higher education release their reopening plans for full on-campus immersion in the 2021 and 2022 academic years, college student attitudes towards taking a COVID-19 vaccine may also change due to idiosyncratic institutional planning. Yet, the strengths of this study is its sample size (n=245), rendering it robust for quantitative analysis and generalizability, while also reporting timely and critical data for institutions of higher education: For these reasons, the research team feels the study’s strengths outweigh its limitations.

## Results

Results from our final sample are presented below. Table 1 below breaks down the descriptive statistics of our nationwide sample.

**Table 1.**
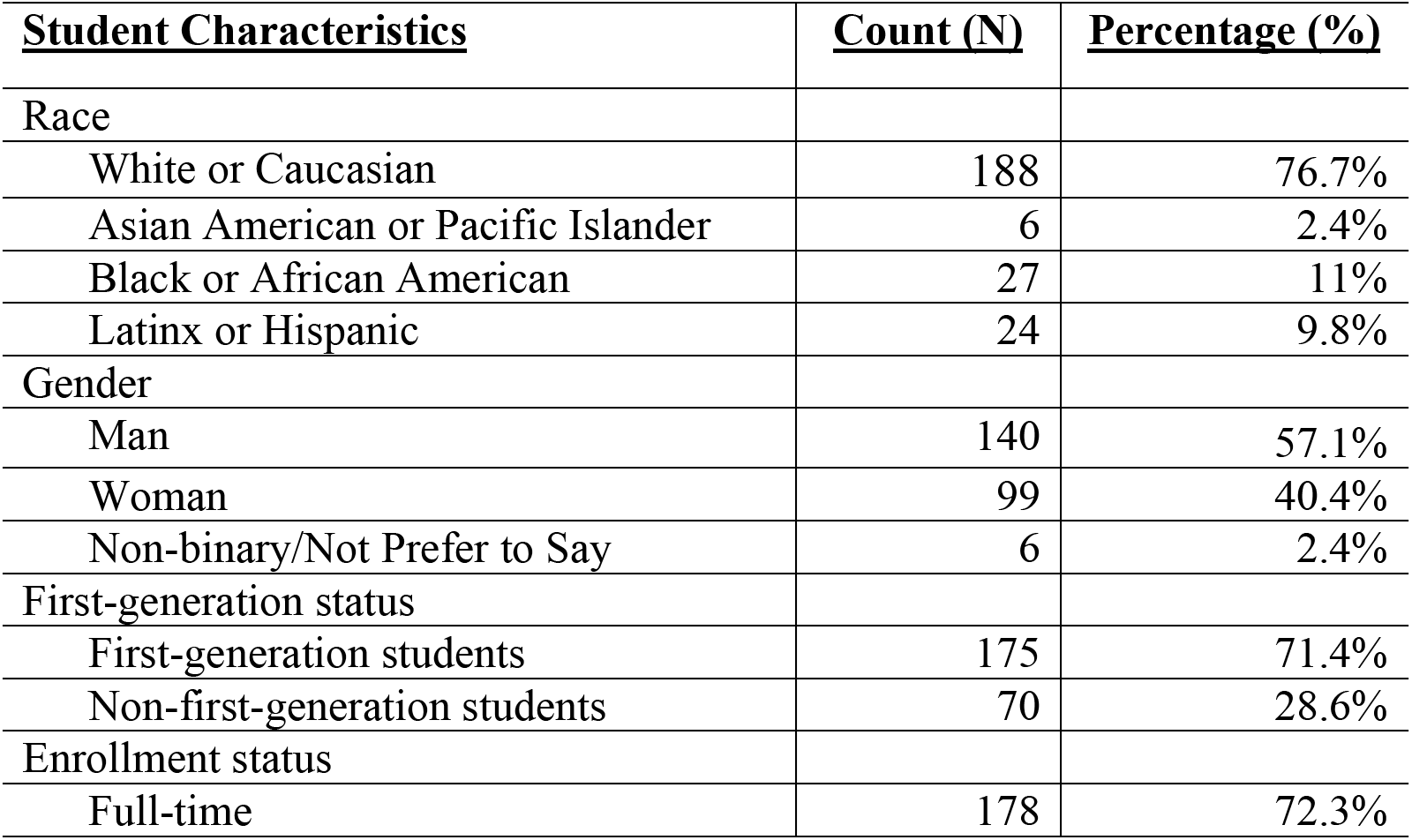
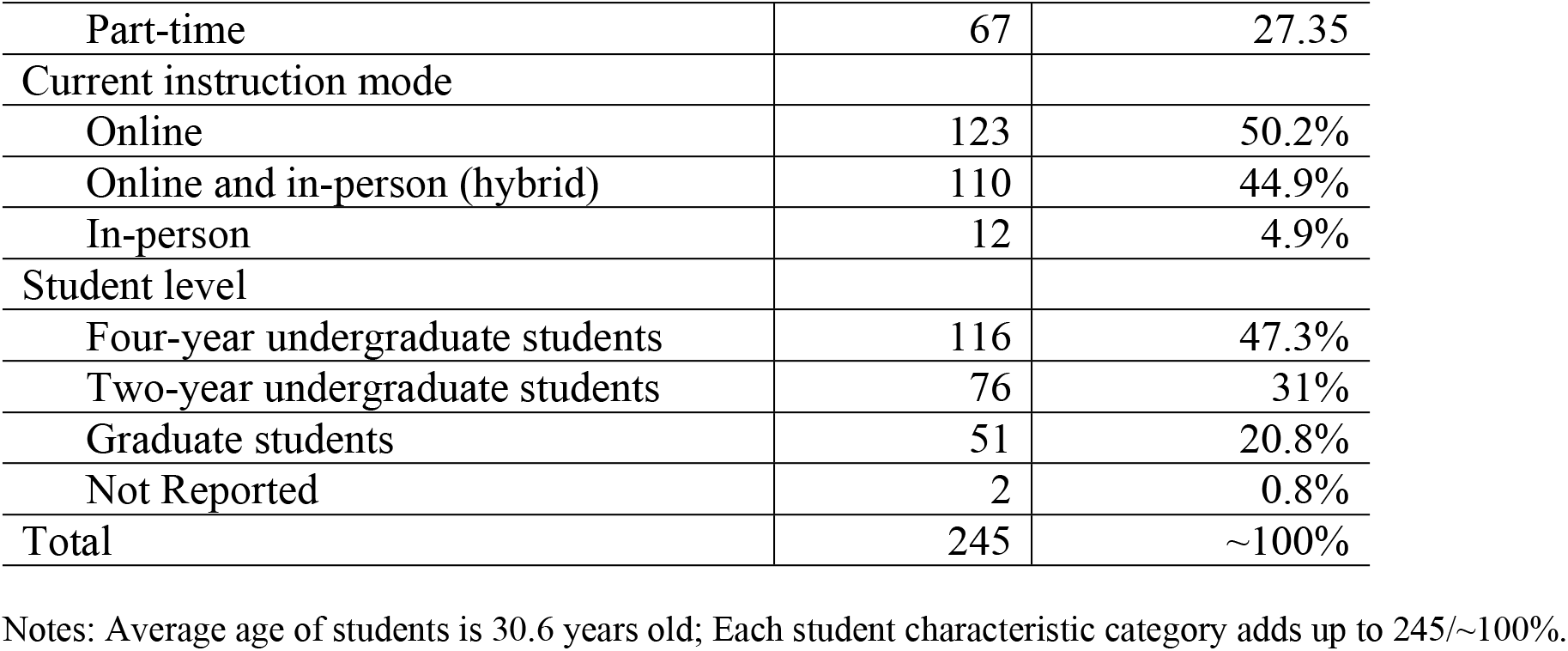
Descriptive statistics of survey sample and responses (N=245)

It should be noted that our sample is relatively representative of the American postsecondary student population given the most recent statistics available from the National Center of Education Statistics (NCES). There were more men respondents than women (57% to 37%, respectively), which is inversely distributed compared to the overall U.S. postsecondary population at large (57.4% women to 42.6% men; De Brey et al., 2021, Table 303.60). Four broad race categories emerged, with the majority of respondents identifying as White or Caucasian (77%), followed by Black or African American (11%), Latinx or Hispanic (9.8%), and Asian American or Pacific Islander (2.4%). The corresponding NCES numbers for these groups are 51.6% White/Caucasian, 7% Asian American/Pacific Islander, 12.7% Black/African American, and 19.2% Latinx/Hispanic respectively (De Brey et al., 2021, Table 306.50). Again, our respondents were relatively representative, with an oversampling of White/Caucasian SWDs and an undersampling of Asian American/Pacific Islander SWDs. The sample did not include any individuals who used the survey’s fill-in-the-blank option to identify as American Indian/Alaska Native, two or more races, or nonresident aliens, with these three groups combined representing approximately 9.5% of the nationwide student population (De Brey et al., 2021, Table 306.50).

The majority of students reported being enrolled in a four-year undergraduate program (47.3%), with 31% enrolled in a two-year undergraduate program, and 20.1% pursuing a postbaccalaureate credential or degree. Four-year undergraduate students were slightly undersampled compared to national enrollment of four-year (55.8%), with two-year (28.5%) programs and graduate students being oversampled (15.6%) (De Brey et al., 2021, Table 303.60). At 72.7%, over two-thirds of respondents were enrolled full-time in their program, with 27.3% reporting part-time status; this full-time-heavy distribution is similar to the at-large population that is divided 61% to 39%, respectively (De Brey et al., 2021, Table 303.60).

The remaining two demographic features, first-generation status and instructional mode, are not collected at the national level by NCES at this time. It should be noted that the vast majority of our sample, 71.4%, identified as first-generation students. Additionally, as expected due to the COVID-19 pandemic and its subsequent impact on academic instruction, many students reported taking their courses either exclusively online (50.2%) or through an online/in-person hybrid (4.9%). Inversely, 44.9% were found to be currently participating in in-person only instruction. THis balance between online students and students taking classes in-person renders this sample a strong one, as many institutions have required COVID-19 vaccines to return to on-campus learning environments, forcing students to choose between online learning without a vaccine or in-person learning with a vaccine (Redden, 2021).

Summary statistics of college students’ with disabilities knowledge of COVID-19 vaccination costs can be found in Table 2 below:

**Table 2.** College students with disabilities knowledge of vaccine costs (N=245)

| **<u>Student Characteristics</u>** | **<u>Free</u>** | **<u>\$1-99</u>** | **<u>\$100-199</u>** | **<u>\$200+</u>** |
| --- | --- | --- | --- | --- |
| Race |  |  |  |  |
| White or Caucasian (n=188) | 56 (22.9%) | 44 (17.9%) | 72 (29.3%) | 16 (6.5%) |
| Asian American or Pacific Islander (n=6) | 0 | 3 (1.2%) | 3 (1.2%) | 0 |
| Black or African American (n=27) | 7 (2.9%) | 6 (2.5%) | 11 (4.4%) | 3 (1.2%) |
| Latinx or Hispanic (n=24) | 3 (1.2%) | 12 (4.9%) | 9 (3.6%) | 0 |
| Gender |  |  |  |  |
| Man (n=140) | 27 (11%) | 41 (16.7%) | 60 (24.5%) | 13 (5.3%) |
| Woman (n=99) | 35 (14.3%) | 23 (9.4%) | 35 (14.3%) | 6 (2.5%) |
| Non-binary/Not Prefer to Say (n=6) | 4 (1.6%) | 2 (0.8%) | 0 | 0 |
| First-generation status |  |  |  |  |
| First-generation students (n=175) | 37 (15.1%) | 52 (21.2%) | 80 (32.7%) | 6 (2.5%) |
| Non-first-generation students (n=70) | 29 (12.2%) | 13 (5.5%) | 15 (6.3%) | 13 (5.5%) |
| Enrollment status |  |  |  |  |
| Full-time (n=178) | 45 (18.4%) | 12 (4.9%) | 78 (31.8%) | 12 (4.9%) |
| Part-time (n=67) | 21 (8.6%) | 22 (9%) | 17 (6.9%) | 7 (2.9%) |
| Current instruction mode |  |  |  |  |
| Online (n=123) | 34 (13.9%) | 41 (16.7%) | 39 (15.9%) | 9 (3.7%) |
| Online and in-person (hybrid) (n=12) | 30 (12.2%) | 17 (6.9%) | 53 (21.6%) | 10 (4.1%) |
| In-person (n=110) | 2 (0.8%) | 7 (2.9%) | 3 (1.2%) | 0 |
| Student level |  |  |  |  |
| Four-year undergraduate students (n=116) | 25 (10.2%) | 40 (16.3%) | 41 (16.7%) | 12 (5.1%) |
| Two-year undergraduate students (n=76) | 23 (9.4%) | 18 (7.3%) | 28 (11.4%) | 7 (2.9%) |
| Graduate students (n=51) | 18 (7.3%) | 7 (2.9%) | 26 (10.6%) | 0 |
| Total | 66 (26.9%) | 65 (26.5%) | 95 (38.8%) | 19 (7.8%) |

Data in Table 2 suggests that only 26.9% of college students with disabilities were aware that the COVID-19 vaccine is free, while the remaining 73.1% believed the COVID-19 vaccine would cost at least $1, with 7.8% of the sample believing that the vaccine would cost $200 or more. There were also differences between groups in terms of race, gender, first-generation in college student status, disability status, enrollment status, instruction mode, and student level.

By race, White/Caucasian students with disabilities were most aware that COVID-19 vaccines are free (22.9%, n=56), while only 2.9% of Black of African American students, 1.2% of Latinx/Hispanic students, and 0% of Asian American/Pacific Islander students were aware that COVID-19 vaccines are free. By gender, women with disabilities were most aware that COVID-19 vaccines are free (14.3%, n=35) with men with disabilities (11%) having comparable knowledge. Although the sample size was small four out of six non-binary conforming students with disabilities indicated that the COVID-19 vaccine is free, the highest percentage of this knowledge of any gender category.

First-generation in college students were more likely to be aware that COVID-19 vaccines are free (15.1%) than peers (12.2%). Moreover, differences were present among students by enrollment status, instruction mode, and level. Full-time students (12 credits or more per semester) were more aware of COVID-19 vaccines being free than their part-time (11 credits or fewer) peers (18.4% versus 8.6%), while students currently learning online were slightly more aware of COVID-19 vaccines being free (13.9%) compared to students learning in hybrid settings (12.2%). Perhaps one of the largest findings of this study was that students with disabilities learning in in-person settings were least likely to understand that COVID-19 vaccines are free (2 of 110 or 0.8%). This is particularly problematic, as students with disabilities learning in-person may be most at-risk of contracting COVID-19 on campus, given the volume of people on campus for a variety of reasons (other students, faculty, staff, administrators, maintenance crews, etc.).

Finally, there were negligible differences between vaccine cost knowledge of students from different institution levels, as 10.2% of students with disabilities attending four-year institutions were aware that COVID-19 vaccines are free compared to 9.4% of students from two-year institutions and 7.3% of students in graduate school. It was somewhat surprising that graduate students with disabilities were least likely to understand that COVID-19 vaccines are free, knowing that these students are likely older and may have more experience with the U.S. healthcare system. Additionally, graduate students may be more avid readers of the news from reputable sources rather than social media, as social media has been found to be mass spreaders of misinformation specifically regarding COVID-19 vaccines and the pandemic more generally (Shearer, 2021). We will address this finding and others in subsequent sections of this study.

### Implications for Policy and Practice, and Conclusion

The U.S. Census Bureau’s March 2021 findings revealed millions of U.S. residents believed they would have to pay—either out of pocket or through insurance—for a COVID-19 vaccine, contributing to vaccine hesitancy of these individuals (United States Census Bureau, 2021). Similarly, this study revealed that many college students with disabilities--potentially half--may believe that the COVID-19 vaccine is not free, and therefore, may be unaffordable. Across the entire sample of 245 responses, only 26.9% of college students with disabilities were aware that COVID-19 vaccines are free. As reviewed in the introduction of this study, there are many different types of vaccine hesitancy which can contribute to the delay of disease containment and the eventual achievement of herd immunity (Salmon et al., 2015). Given the U.S. Census Bureau’s national study and the college student study at hand, both COVID-19 providers and institutions of higher education must provide clear health communication to both current and prospective students about the costs of the COVID-19 vaccine. Given the data in this study, it was clear that some groups of students with disabilities knew much more than others about COVID-19 vaccine costs, which may contribute to vaccine hesitancy.

First, and perhaps one of the most critical findings of this study, was that White/Caucasian students with disabilities were much more aware of COVID-19 vaccines being free than students with disabilities of Color, possibly further minoritizing these students and increasing their vaccine hesitancy. Decades of research has documented the fact that communities of Color in general are more skeptical about the U.S. healthcare system given historical atrocities such as the Tuskegee Experiment (Suite et al., 2007). As a result, institutions of higher education--and the U.S. healthcare system--must do more to communicate with students of Color with disabilities to ensure that they are being provided with accurate information and have every opportunity to access a COVID-19 vaccine if they so choose.

Next, students with disabilities attending classes in-person were least likely to be aware that COVID-19 vaccines are free--this is a particularly troubling finding, as many institutions have touted their vaccine communication efforts and on-campus vaccination programs (Redden, 2021). However, data in this study suggest that students with disabilities--although in-person and on-campus--may not be receiving accurate information or adequate information regarding COVID-19 vaccine costs. Given that students with disabilities learning in-person may be most at-risk of contracting COVID-19 on campus, institutions of higher education and their disability services offices or student health offices must do more to communicate COVID-19 related policies, including cost structures and vaccine availability.

Finally, and also surprising, was that data in this study suggested that graduate students with disabilities were least likely to be aware that COVID-19 vaccines are free despite graduate school aged students likely being older and having more experience with the U.S. healthcare system--or vaccines--than younger students with disabilities attending two- and four-year institutions of higher education. Although the knowledge of COVID-19 vaccine costs were overall low in this study (26.9% of all students with disabilities knew COVID-19 vaccines were free), it is perplexing that graduate students with disabilities were not as knowledgeable as their peers regarding vaccine costs.

As previously mentioned, older people in the U.S. tend to seek out news from reputable news outlets rather than anonymous social media sources (Shearer, 2021), and these news seeking behaviors would lead one to believe that graduate students--a likely older population of students--would follow this trend and perhaps be more knowledgeable of COVID-19 vaccine costs. However, emerging research has found that digital communication of vaccine policies has become increasingly difficult given the sheer volume of online news sources available and the rise of fake news and misinformation on many online platforms, including social media (Thomas & Pollard, 2020). From here, institutions of higher education and healthcare systems need to engage with social media platforms and other sources of media regularly consumed by college students to properly communicate COVID-19 vaccine policies, including vaccine costs. Moreover, targeted communication should specifically address college students with disabilities to alert them to vaccine policies and costs. This effort can start with disability services offices contacting all known students with disabilities by phone, even if this type of campaign is costly in terms of time and human resources--the information is critical and institutions must step up to ensure that one of their most vulnerable populations has all of the information necessary to make informed decisions regarding their own health, the health of others, and the health of their entire campus community.

## Discussion

There has been a plethora of pre-COVID-19 research that has shown the need for accurate, updated information on vaccine efforts to the general public by organizations like the U.S. federal government (Eskola et al., 2015). In summer 2020, although the Centers for Disease Control and Prevention (CDC), along with the U.S. federal government, stated that the costs of COVID-19 vaccines would be financed by emergency funds and that U.S. residents, including persons with disabilities, could get a COVID-19 vaccine for free (Centers for Disease Control and Prevention, 2021), this message does not appear to be disseminated widely after one year into the pandemic, as false information regarding vaccine costs after the CDC made its announcement have been circulated (Sauer et al., 2021). As a result, persons may have overlooked or not remembered that COVID-19 vaccines are free for U.S. residents, or there is a possibility that these individuals may have developed vaccine hesitancy due to misinformation surrounding the cost of these vaccines.

Throughout the past year, the hesitancy to receive the vaccine based on cost can potentially be detrimental to college students with disabilities as well as higher education institutions. In this study, less than 25% of students with disabilities believed that the COVID-19 vaccines were free. It can be argued that the student disability population is the most at-risk for COVID-19 effects and vaccine effects. In these instances, students with disabilities may already have some degree of Salmon et al.’s (2013) *manmade hesitancy*, and these students may also believe these COVID-19 vaccines are unaffordable. As this study highlighted, White/Caucasian students with disabilities were more informed of the vaccine being free, versus those students with disabilities from other racial/ethnic backgrounds. It can be argued that students with disabilities of color may already feel minoritized by higher education institutions thus increasing their vaccine hesitancy. To target this informational discrepancy between White/Caucasian students with disabilities and students with disabilities of color, it is necessary for both governmental and higher education organizations to provide credible and scientific information on COVID-19 vaccine costs, side effects, and general information. Working with organizations in the community and local agencies could assist in both vaccination and information access efforts for students with disabilities within higher education institutions.

According to Jarrett et al. (2015) raising awareness and knowledge through credible medical sources has been instrumental in lowering vaccine hesitancy for years. However, it is important that this information is shared consistently on a continuous basis and attempt to reach the wider public through public addresses by government officials, prioritizing the location of these guidelines on the CDC and U.S. federal government websites via multiple modes of digital communication such as text messaging, email, social media, and physical communication, in addition to word-of-mouth from health-care workers (Jarrett et al., 2015). In large, urban areas during the beginning of the COVID-19 pandemic, the vaccination campaign in the United States were used as mass vaccination sites, and as such higher education institutions should also communicate this information clearly (Redden, 2018). However, after thorough review of the communication from several higher education institutions, the researchers of the study indicated that vaccine cost was not communicated. Interestingly, one researcher on the team was asked to bring their health insurance card although the CDC stated that this was not a requirement, as these vaccines are free regardless of insurance status.

Additionally, the CDC’s 2021 announcement that the COVID-19 vaccines would be free there was no indication of vaccine cost and the vaccine being free in the list of “Key Things to Know”(Centers for Disease Control and Prevention, 2021, para. 1). In fact, there was only mention of making the COVID-19 vaccines more widely accessible (para. 5) without any other mention of cost in the main menu. The CDC website only mentioned the “Cost of Vaccines” (para. 31) in the 33rd paragraph and indicated that the”federal government is providing the vaccine free of charge to all people living in the United States, regardless of their immigration or health insurance status” (Centers for Disease Control and Prevention, 2021, para. 33). It is important that organizations like the CDC and higher education institutions should prioritize that vaccines are available free of charge and continue to raise awareness in this regard to potentially lower vaccine hesitancy among students with disabilities.

Furthermore, information and communication regarding COVID-19 vaccines could consider not mentioning information about health insurance as it may be misleading that the COVID-19 vaccine is not free and that insurance is needed. Messages from the government, healthcare, and educational institutions could simply convey the messages that COVID-19 vaccines are safe, effective and free. By simplifying this message, there may be a decrease in vaccine hesitancy among many groups of U.S. residents, including college students with disabilities. For higher education institutions seeking to re-open campuses safely via COVID-19 requirements, their messaging regarding the vaccine should be clear that the vaccine is free, and this should be communicated clearly and frequently on all platforms to reach as many students as possible. In fact, students with disabilities should receive clear communication regarding COVID-19 vaccines from all Student Support Services offices on campus. While the literature has shown that students may experience any of the vaccine hesitancies as highlighted Salmon et al (2015), but COVID-19 vaccine hesitancy specifically with regards to cost can be resolved by clear and effective communication to the students.

Ultimately, higher education institutions should not assume that their students understand the COVID-19 policies outlined by the federal government as seen by the significant number of students with disabilities who were unaware that the COVID-19 vaccine is free. For college students with disabilities to continue with their goals towards graduation and for higher education institutions to remain economically stable, it is imperative that COVID-19 vaccine policies and practices are communicated clearly and efficiently so that cost-related vaccine hesitancy will not contribute to the detriment of U.S. society’s public health and the education of its citizens.

## Data Availability

All data produced in the present study are available upon reasonable request to the authors

## Notes

### Competing Interest Statement

The authors have declared no competing interest.

### Funding Statement

This study did not receive any funding

### Author Declarations

The institutional review board of Texas State University gave ethical approval for this work.

